# The urgent need for phased university reopenings to mitigate the spread of COVID-19 and conserve institutional resources: A modeling study

**DOI:** 10.1101/2020.08.25.20182030

**Authors:** Lior Rennert, Corey A. Kalbaugh, Christopher McMahan, Lu Shi, Christopher C. Colenda

**Affiliations:** Department of Public Health Sciences, Clemson University, Clemson, SC, USA; School of Mathematical and Statistical Sciences, Clemson University, Clemson, SC, USA; Principal, C.C. Colenda, LLC; Section of Gerontology and Geriatrics, Department of Internal Medicine, Wake Forest University, NC, USA

## Abstract

**Introduction:** Recent outbreaks of COVID-19 in universities across the United States highlight the difficulties in containing the spread of COVID-19 on college campuses. While research has shown that mitigation strategies such as frequent student testing, contact tracing, and isolation of confirmed and suspected cases can detect early outbreaks, such mitigation strategies may have limited effectiveness if large outbreaks occur. A phased reopening is a practical intervention to limit early outbreaks, conserve institutional resources, and ensure proper safety protocols are in place before the return of additional students to campus.

**Methods:** We develop dynamic compartmental transmission models of SARS-CoV-2 to assess the impact of a phased reopening and pre-arrival testing on minimizing outbreaks (measured by daily infections) and conserving university resources (measured by isolation bed capacity). We assume that one-third of the student population returns to campus each month as part of the phased reopening, and that pre-arrival testing removes 90% of infections at the semester start. We assume an on-campus population of *N* = 7500, an active COVID-19 prevalence of 2% at baseline, and that 60% of infected students require isolation for an average period of 11 days. We vary the reproductive number (*R_t_*) between 1.25 and 4 to represent the effectiveness of alternative mitigation strategies throughout the semester, where *R_t_* is constant or improving throughout the semester (ranging from 4 to 1.25).

**Results:** Compared to pre-arrival testing only or neither intervention, phased reopening with pre-arrival testing reduced peak daily infections by 6% and 18% (*R_t_*=1.25), 44% and 48% (*R_t_*=2.5), 63% and 64% (*R_t_*=4), and 72% and 74% (improving *R_t_*), respectively, and reduced the proportion of on-campus beds needed for isolation from 10%-25% to 5%-9% across different values of *R_t_*.

**Conclusion:** Phased reopening with pre-arrival testing substantially reduces the peak number of daily infections throughout the semester and conserves university resources compared to strategies involving the simultaneous return of all students to campus. Phased reopenings allow institutions to improve safety protocols, adjust for factors that drive outbreaks, and if needed, preemptively move online before the return of additional students to campus, thus preventing unnecessary harm to students, institutional faculty and staff, and local communities.

## Introduction

Higher education institutions are struggling to reopen their campuses in a safe and judicious manner during the COVID-19 pandemic. The reopening strategies for Fall 2020 implemented by several universities in the United States have largely failed.^1,2^ Other institutions have elected to delay reopening, partially reopen, or remain closed, preferring instead to continue with online instruction.^3-5^ Those planning to continue with reopening are exploring several preventative strategies to mitigate the spread of COVID-19, including frequent SARS-CoV-2 testing, contact tracing, and isolation of confirmed and suspected cases.^3,4^

Previous modeling studies have demonstrated that high numbers of active infections at the beginning of the semester lead to early and large outbreaks and drain institutional resources.^5^ This has been further evidenced by recent COVID-19 outbreaks in universities, which have been forced to send students home and shift to online learning within one week of reopening.^1,2^ With such early outbreaks and closures, implementation of preventative strategies throughout the semester may no longer be relevant, as these strategies are intended to prevent outbreaks rather than contain them. Indeed, the initial number of active infections assumed by modeling studies that support these strategies^3,6,7^ may be far lower than suggested by current estimates of SARS-CoV-2 prevalence^5,8^ and recent university reopenings.^1,2^ Colleges and universities intending to reopen campuses in Fall 2020 or in future semesters must therefore place a greater emphasis on reducing active infections and limiting outbreaks at the semester start. However, given the number of high-density social gatherings that occur early in the semester and beyond,^1,9,10^ such outbreaks may be difficult to contain with the simultaneous return of all students to campus.^11^

A phased return of students to campus is a practical intervention to limit early outbreaks, ensure proper protocols are in place, and conserve university resources. This is accomplished through minimizing the susceptible population size and density early in the semester, which can delay large outbreaks and reduce outbreak size, ensure the availability of sufficient resources by vacating a large portion of isolation beds for confirmed or suspected cases, and increase testing and support service capacity per student. Furthermore, a phased reopening provides institutions time to improve strategies to mitigate the spread of COVID-19 and adjust for factors that drive outbreaks (e.g., fraternity gatherings^1,9,10^) before the return of additional students to campus. If outbreaks cannot be contained, a phased reopening allows higher education institutions to pre-emptively transition to remote learning before the arrival of all students and thus prevent unneccessary harm.

Our team was tasked with recommending strategies to limit outbreaks and ensure adequate resources for confirmed on-campus COVID-19 cases in a large university in the Southeastern United States. To guide and inform our recommendations, we developed dynamic compartmental transmission models to assess the impact of a phased reopening, along with exclusion of COVID-19 positive students through testing prior to campus arrival, on minimizing outbreak size and conserving university resources throughout the semester.

## Methods

To capture the essential features of COVID-19 spread on campus, we developed dynamic compartmental transmission models of SARS-COV-2^12^ with the following compartments: susceptible, exposed, infectious (asymptomatic), infectious (symptomatic), isolated, and recovered (**Figure 1**). We assumed a large on-campus population (*N* = 7,500), an active infection rate of 2% at the semester start,^13^ and that 60% of active infections would be detected and require isolation^14^ for an average period of 11 days.^15^ We considered four settings for the reproductive number (*R_t_*) to represent the effectiveness of various mitigation strategies throughout the semester: highly effective (*R_t_* = 125), moderately effective (*R_t_* = 2.5), and ineffective (*R_t_* = 4), along with a time-varying *R_t_* that improved throughout the semester (*R*_0_ = 4, *R_t_* = 2.5, and *R_t_* = 1.25 for months *t* ≥ 2). The latter setting was intended to capture improvement in mitigation efforts over time. Additional assumptions were compiled from published sources and are provided in **Table 1**. Because this study used only a theoretical model with no actual data, the Institutional Review Board of Clemson University determined that this research did not involve human participants and did not require their approval.

**Figure 1.**
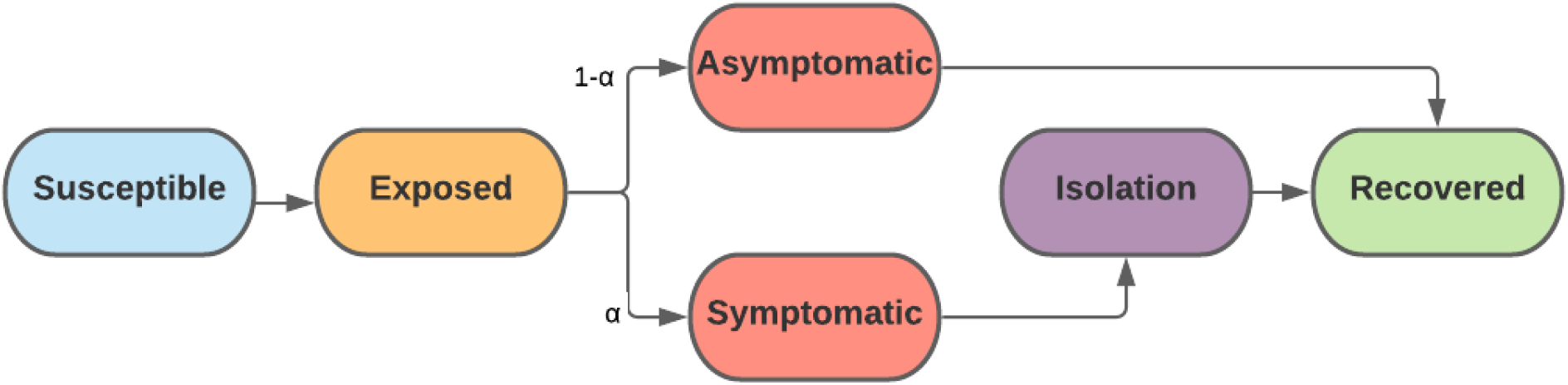
Model structure. The population is divided into the following six compartments: susceptible, exposed (not infectious or detectable), asymptomatic (infectious and not detectable), symptomatic (infectious and detectable), isolated (infectious but no contact with other individuals), and recovered. Exposed individuals transition into the symptomatic compartment with probability α and asymptomatic compartment with probability 1-α. It is assumed all symptomatic individuals are detected (after an average period of 2 days) and isolated for the remaining duration of their infection.

**Table 1:** Model input 112 parameters, assumptions, and references.

| Model parameter | Input |
| --- | --- |
| On-campus population (N) | 7,500 (assumption) |
| ...Other compartments | 0 (assumption) |
| Time horizon (months) | 3 months<sup>7</sup> |
| Disease dynamics<sup>a</sup> |  |
| ...Mean incubation time, $1/\sigma$ | 3 days<sup>17</sup> |
| ...Mean asymptomatic infectious time (days), $1/\phi$ | 14 days<sup>17</sup> |
| ...Mean symptomatic infection time before detection and isolation (days), $1/\gamma$ | 2 days<sup>17</sup> |
| ...Isolation time, $1/\rho$ (days) | 11 days: Time between symptom onset and negative NAT test<sup>15</sup> |
| ...Proportion of infections that are symptomatic, $\alpha$ | 0.6<sup>14</sup> |
| Transmission rate, $\beta$ | Dependent on $R_t$ |
| Baseline infectious rate (%)<sup>b</sup> | 2%<sup>13</sup> |
| Baseline recovered rate (%) | 10%<sup>5,7,8</sup> |
| Mitigation strategies throughout semester ($R_t$)<sup>c</sup> | |
| ...Highly effective (best case) | 1.25<sup>18</sup> |
| ...Moderately effective (base case) | 2.5<sup>3,14</sup> |
| ...Ineffective (worst case) | 4<sup>14</sup> |
| ...Time-varying | Month 0: 4; Month 1: 2.5; Month 2+: 1.25 (assumption) |
| Interventions |  |
| Test characteristics |  |
| ...Sensitivity (%) | 90%<sup>3,19</sup> |
| ...Specificity (%) | 100% (assumption) |
| Phased re-opening |  |
| ...Phase 1: Calendar time (months)/sub-population returning to campus | 0 months/2,500 students (assumption) |
| ...Phase 2: Calendar time (months)/sub-population returning to campus/cumulative population | 1 months/2,500 students/5,000 students (assumption) |
| ...Phase 3: Calendar time (months)/sub-population returning to campus/cumulative population | 2 months/2,500 students/7,500 students (assumption) |

<sup>a</sup> We assume a closed system (i.e., no exogenous infections or deaths)
<sup>b</sup> Under pre-arrival testing, baseline infections are reduced by 90%
<sup>c</sup> Under phased reopening, we assume this number is reduced by 20% during the first phase and 10% during the second phase

We considered three interventions: Phased reopening with pre-arrival testing, pre-arrival testing only, and neither intervention. We assumed that infectious students enter campus through the asymptomatic compartment, and that pre-arrival testing reduced the number incoming infections by 90%. We assumed a phased re-opening over a 2-month period, in which one-third of the population (*N\** = 2500) returns to campus at the semester start, 30 days after the semester start, and 60 days after the semester start. We further assumed that the reproductive rate is reduced by 20% during the first phase and 10% during the second phase due to a decrease in student population density, and that any unoccupied beds during these phases are available for isolation of detected COVID-19 cases. The equations and initial values for each compartment are provided in **Supplementary Table 1**.

We evaluated the relative impact of each intervention on the number of new active infections throughout the semester and isolation bed capacity. At each timepoint *t*, new active infections were defined as entries into the asymptomatic infectious and symptomatic infectious compartments. Isolation bed capacity was measured as the number (*n_beds_*) and proportion (*n_beds_/N*) of on-campus beds needed for isolation throughout the semester. We also evaluated the impact of each strategy on the time until 5% of on-campus beds were occupied for isolation of confirmed cases. The latter metric is based on recommendations to reserve enough isolation beds for 5% of the on-campus student population,^20^ which is currently higher than the isolation bed capacity of at least several large US universities.^21,22^ With an on-campus population of 7,500, a threshold of 5% would require reservation of 375 beds for isolation.

## Results

The number of new (active) infections throughout the semester based on the model projections are displayed in **Figure 2**. In all scenarios, both pre-arrival testing alone and in conjunction with phased reopening reduced the rate of new daily infections early in the semester and delayed the timing of the peak outbreak size. A phased reopening in conjunction with pre-arrival testing reduced the size of the peak outbreak across all scenarios. Under highly effective mitigation strategies throughout the semester (*R_t_* = 1.25), a phased reopening reduced the peak by 6% and 18% compared to pre-arrival testing only and no interventions, respectively. For moderate to ineffective mitigation strategies throughout the semester, this decrease was substantial. Compared to the simultaneous return of all students, a phased reopening (with pre-arrival testing) decreased the peak number of new infections between 44% to 48% (*R_t_* = 2.5) and 63% to 64% (*R_t_* = 4). Under improving mitigation strategies throughout the semester (*R*_0_ = 4, *R_t_* = 2.5, and *R_t_* = 1.25 for *t ≥* 2 months), phased reopening with pre-arrival testing decreased the peak number of new infections between 72% to 74%. Meanwhile, improvement in mitigation efforts throughout the semester had little impact on reducing the rate of new infections for strategies involving the simultaneous return of students to campus. Compared to no interventions, pre-arrival testing alone reduced the peak number of new infections between 6% and 12% across all scenarios.

Pre-arrival testing alone had minimal impact on maximum isolation bed capacity, while implementation in conjunction with a phased reopening substantially reduced the number of isolation beds needed for values of *R_t_* > 2.5 (**Table 2**). In these scenarios, phased reopening with pre-arrival testing required reservation of 7% of on-campus beds for isolation, compared to 20% (*R_t_* = 2.5) and 25% (*R_t_* = 4) for the other interventions. Under improving *R_t_*, phased reopening in conjunction with pre-arrival testing required 5% of on-campus beds for isolation. With highly effective mitigation strategies throughout the semester (*R_t_* = 1.25), there was little difference in the maximum number of isolation beds needed (range: 9% to 11%).

**Table 2.**
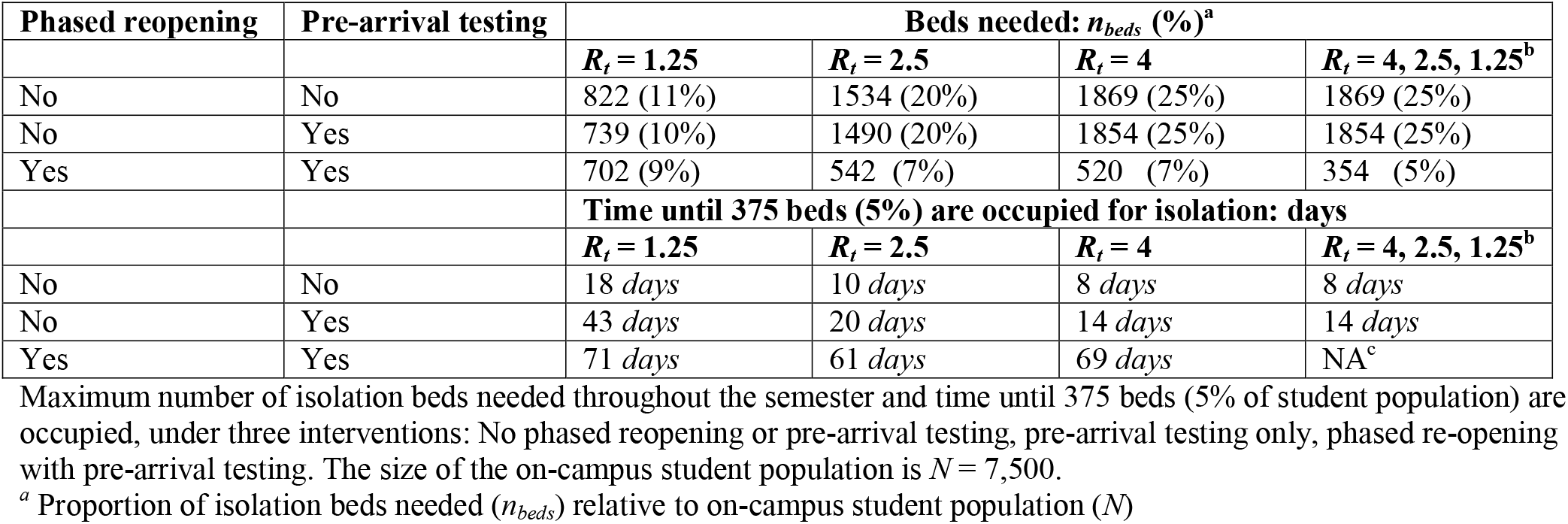
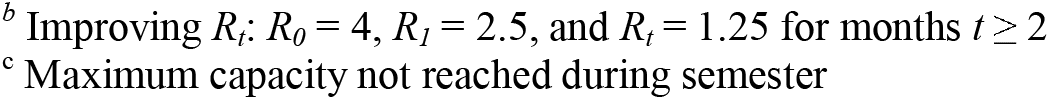
Modeling results for occupied isolation beds for each strategy across varying disease reproductive numbers.

| Phased reopening | Pre-arrival testing | Beds needed: $n_{beds}$ (%)<sup>a</sup> | | | |
| --- | --- | --- | --- | --- | --- |
| | | $R_t = 1.25$ | $R_t = 2.5$ | $R_t = 4$ | $R_t = 4, 2.5, 1.25^b$ |
| No | No | 822 (11%) | 1534 (20%) | 1869 (25%) | 1869 (25%) |
| No | Yes | 739 (10%) | 1490 (20%) | 1854 (25%) | 1854 (25%) |
| Yes | Yes | 702 (9%) | 542 (7%) | 520 (7%) | 354 (5%) |
|  |  | Time until 375 beds (5%) are occupied for isolation: days |  |  |  |
| | | $R_t = 1.25$ | $R_t = 2.5$ | $R_t = 4$ | $R_t = 4, 2.5, 1.25^b$ |
| No | No | 18 days | 10 days | 8 days | 8 days |
| No | Yes | 43 days | 20 days | 14 days | 14 days |
| Yes | Yes | 71 days | 61 days | 69 days | NA<sup>c</sup> |

Maximum number of isolation beds needed throughout the semester and time until 375 beds (5% of student population) are occupied, under three interventions: No phased reopening or pre-arrival testing, pre-arrival testing only, phased re-opening with pre-arrival testing. The size of the on-campus student population is $N = 7,500$ .
<sup>a</sup> Proportion of isolation beds needed ( $n_{beds}$ ) relative to on-campus student population ( $N$ )
<sup>b</sup> Improving $R_t$ : $R_0 = 4$ , $R_t = 2.5$ , and $R_t = 1.25$ for months $t \geq 2$
<sup>c</sup> Maximum capacity not reached during semester

Reservation of 375 beds for isolation (5% of on-campus student population) was not sufficient without a phased reopening. Under no interventions or pre-arrival testing only, this threshold was reached in 18 and 43 days for *R_t_* = 1.25, 10 and 20 days for *R_t_* = 2.5, and 8 and 14 days for *R_t_* = 4, respectively. A phased reopening substantially increased the time until this threshold was reached under these scenarios (range: 61 to 71 days). This intervention under improving mitigation efforts throughout the semester (improving *R_t_*) was the only scenario in which isolation bed occupancy did not exceed the 5% threshold.

**Figure 2:**
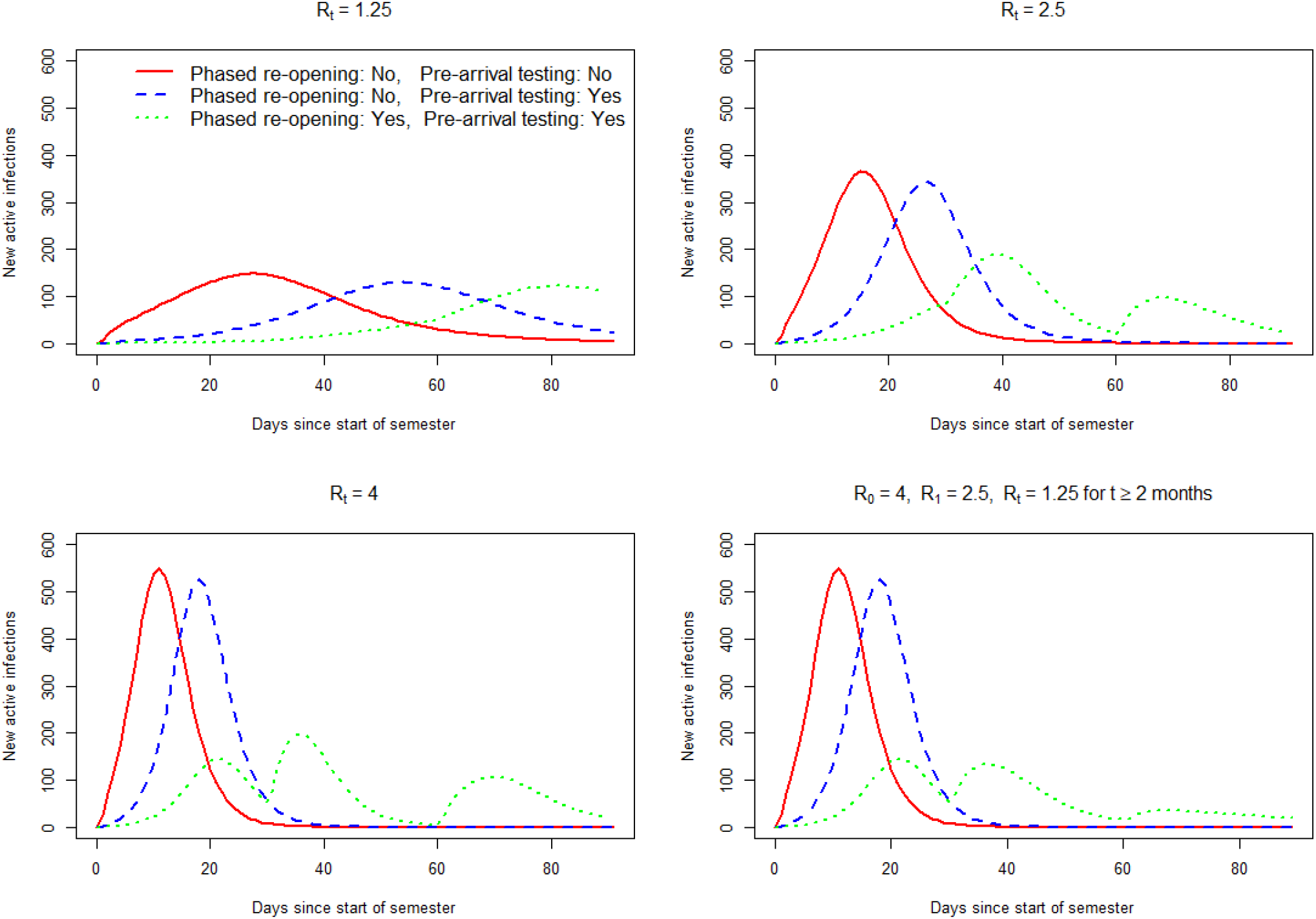
Projected new infections (daily) throughout the semester for each intervention. Expected number of new active infections throughout the semester under three interventions: No phased reopening or pre-arrival testing (solid red line), no phased reopening with pre-arrival testing (dashed blue line), phased reopening with pre-arrival testing (dotted green line). Top left panel: effective mitigation strategies throughout semester (*R_t_* = 1.25). Top right panel: moderately effective mitigation strategies throughout the semester (*R_t_* = 2.5). Bottom left panel: ineffective mitigation strategies throughout the semester (*R_t_* = 4). Bottom right panel: improving mitigation strategies throughout the semester (*R_0_* = 4, *R_1_* = 2.5, and *R_t_* = 1.25 for months *t≥* 2). New active infections are defined as new entries into the asymptomatic infectious and symptomatic infectious compartments.

## Discussion

Minimizing the number of active infections at the start of the semester is essential to limiting rapid outbreaks and ensuring available resources for support services such as testing, contact tracing, and case isolation. We found that unless highly effective mitigation strategies are implemented throughout the semester, which may be optimistic given recent events,^1,2^ the simultaneous arrival of all students to campus leads to early and large outbreaks. Furthermore, the timing of these outbreaks occur early in the semester and are therefore not impacted by improvements in mitigation efforts over time. Alternatively, a phased return of students to campus in conjunction with testing prior to arrival substantially delays the peak outbreak timing and reduces the outbreak size by up to 74%.

Rapid outbreaks drain university resources and will inevitably lead to institutions shutting down on-campus activities and shifting fully online. Our models demonstrate that the simultaneous return of all students to campus may require 10% to 25% of on-campus beds for isolation of confirmed COVID-19 cases, far exceeding the current capacity of several large institutions.^21,22^ In fact, we found that the recommended 5% of isolation beds for confirmed COVID-19 cases^20^ may lead to institutions reaching capacity in less than 10 days. Under a phased reopening with pre-arrival testing, the number of isolation beds needed ranged from 5% to 9% of total capacity.

While previous studies have concluded that frequent SARS-CoV-2 testing and contract tracing can significantly reduce disease spread, they have limitations in college and university settings. First, the models that support these conclusions assume a low number of initial active infections. Contact tracing, for example, has shown to be ineffective when the number of initial infections is greater than 40.^6^ Other modeling studies demonstrating the effectiveness of frequent testing for SARS-CoV-2 assume that initial infections range between 1 and 10 individuals.^3,7^ These are much lower than the number of early infections from recent university outbreaks^1,2^ and current prevalence estimates of SARS-CoV-2.^5,8^ Second, the logistical contraints and high costs may challenge the ability of institutions to impelement these strategies effectively. This may be especially difficult at the semester start, where larger outbreaks are more likely to occur due to the number of high-density social events in these settings.^1,9,10^ Such gatherings have been the driving factor behind several university closings.^1,2^

## Limitations

Our study has several limitations. First, we omit an in-depth discussion of the logistics behind a phased reopening. Difficulties in implementation include the careful coordination of the return of students back to campus and may require institutions to shift between on-campus and remote learning throughout the phased reopening. The costs of unutilized institutional facilities are also not considered here. In addition, our study does not consider the contribution of off-campus students to disease spread, nor does it consider their impact on institutional resources.

While we used current evidence to inform plausible biological parameters for SARS-CoV-2, these values may need refinement as more data become available. However, we performed sensitivity analyses under several disease transmission parameters ranging from best-case to worst-case scenarios in order to provide a range of plausible estimates. Finally, while the effects of mitigation strategies throughout the semester, such as frequent testing, successful contact tracing, and quarantine of suspected cases are implicitly incorporated into our model through the reproductive number (*R_t_*), we do not consider the impact of these strategies on the occupation of reserved beds. Therefore the number of beds needed for reservation may far exceed estimates provided here.

## Conclusions

Based on our findings, we encourge institutions to immediately increase the number of isolation beds and support service capacity. In addition, all students should be tested prior to campus arrival to avoid the return of infectious students to campus. A phased reopening offers several additional benefits. Limiting the number of students on-campus at the start of the semester can substantially reduce the number of initial infections and delay the timing and size of outbreaks, while providing opportunities to improve safety protocols and adjust for factors that drive these outbreaks before returning additional students to campus. Furthermore, minimizing the size of the susceptible population will help ensure that institutions have sufficient resources at their disposal to handle early outbreaks. While mitigation strategies are promising tools to prevent rapid disease spread,^3,6,7^ the logistics of implementation during large outbreaks and their effectiveness in containing them have yet to be tested. A phased reopening provides opportunities to trial such strategies on a smaller population before they are implemented in larger scales. Most importantly, if COVID-19 outbreaks cannot be kept under control with a limited student population, a phased reopening provides the ability to halt the return of additional students to campus and will undoubtedly prevent hundreds to thousands of additional infections, thus preventing unnecessary harm to students, institutional faculty and staff, and local communities.

## Data Availability

No data was collected for this study. R code for dynamic compartmental transmission models will be posted on github, or can be accessed by emailing corresponding author.

## Author contribution statement

LR: Conceptualization, literature search, figures, methodology, writing, review, and editing; CAK: Project administration, writing, review, and editing; CM: Methodology, review, and editing; LS: Review and editing, literature search; CCC: Conceptualization, literature search, writing, review, and editing.

## Supplement

Equations and initial values for dynamic compartmental transmission models

